# The association between dietary diversity and depressive symptoms among adult population in rural western Kenya

**DOI:** 10.64898/2026.03.12.26348250

**Authors:** Charles Opondo, James Kamadi, James Akiruga Amisi, Sonak Pastakia, Molly Rosenberg

**Affiliations:** Department of Epidemiology and Biostatistics, Indiana University School of Public Health, 1025 E. 7TH Street, Bloomington, Indiana, USA; Academic Model Providing Access to Healthcare (AMPATH), P.O. Box 4606, 30100,Eldoret, Kenya; Department of Family Medicine, Moi University School of Medicine, P.O. Box 4606 30100, Eldoret, Kenya; Purdue Kenya Partnership, Purdue University College of Pharmacy, 640 Eskenazi Ave. Indianapolis, IN 46202,USA

**Author notes:** **Corresponding Author:** Charles Opondo, Department of Epidemiology and Biostatistics. Indiana University School of Public Health, 1025 E. 7TH Street, Bloomington, Indiana, USA.

**Keywords:** Dietary diversity, Depressive symptoms, depression, Rural western Kenya

## Abstract

**Introduction:** Low dietary diversity has been identified as a predictor of depression outcomes in high-income countries, while evidence is scarce from low-income settings where poor nutrition and depression often co-occur. In this study, we estimate the relationship between dietary diversity and depression among adults in rural western Kenya.

**Method:** We conducted a cross-sectional analysis of 311 participants enrolled in the Bridging Income Generation through Group Integrated Care program. We assessed depressive symptoms using the 20-item Centre for Epidemiologic Studies for Depression Scale (CES-D), and measured dietary diversity as the number of five food groups consumed in previous 24-hours using a validated dietary diversity scale. We used linear regression to estimate the association between high dietary diversity (consumption of all the five food groups) and continuous depression scores, adjusting for key covariates. We tested for effect measure modification by wealth status. We conducted a secondary analysis using quantile regression to explore variation across depression scores distributions.

**Results:** Higher dietary diversity was associated with fewer depressive symptoms [adjusted β (95% CI): -3.49 (-6.62, -0.38)]. The association was stronger among individuals with low wealth backgrounds [adjusted β (95% CI): -6.00 (-10.46, -1.42)] relative to those with high wealth backgrounds [adjusted β (95% CI): -0.53 (-4.76, 3.68); Wald p-value for interaction term =0.0003]. The effect sizes for the association were larger at higher quantiles, notably at 75th [adjusted β (95% CI): -4.00 (-10.13, 2.13)] and 90th [adjusted β (95% CI): -1.59 (-7.43, 4.25)] compared to those at lower quantiles for 10^th^ [adjusted β (95% CI): -0.59 (-2.46, 1.28) and 25^th^ [adjusted β (95% CI): -0.82 (-4.14, 2.50), though wide confidence intervals limited the precision of effect estimates.

**Conclusion:** In rural western Kenya, higher dietary diversity was associated with lower depression symptoms, particularly among participants from lower wealth backgrounds, and particularly among those with scores consistent with more severe depression symptoms. These findings suggest that improving dietary diversity may offer mental health benefit to the most socioeconomically disadvantaged individuals and could be a promising strategy to reduce depression in resource poor settings. Future work could leverage longitudinal and experimental studies for improved inference and should investigate mechanisms through which dietary diversity may influence depression.

## Introduction

Depression is a major public health concern globally. Approximately 332 million people worldwide experience depressive symptoms (1). In Kenya, the burden of depression is high at 1.9 million cases, placing it among the top five countries for depression in sub-Saharan Africa (2). Further, nearly 25% and 40% of outpatient and inpatients respectively suffer from depression, followed by stress and anxiety disorders (3). In 2019, depression accounted for 33% of the total Disability Adjusted Life Years (DALYs) among adults (4). Although the specific aetiology of depression is unknown, current evidence attributes the occurrence of depression to a complex interaction of social (5), genetic, environmental (6), and biological factors (7). Given the burden that depression places on both individuals’ quality of life and the public health system (8,9), it is important to examine how modifiable lifestyle factors such as diet shape depression outcomes. This need is especially critical in settings where undernutrition and depression frequently co-occur.

An increasing body of evidence suggest that diet may be utilized as part of a prevention strategy for depression. Diet affects various physiological processes that may play a role in the onset of depression, such as inflammation, oxidative stress, or hormonal influences (10). However, compared to other non-communicable diseases, the role of diet in the prevention of depression is still a relatively new field of research (11), particularly in low-income settings. Moreover, prior research on the effect of diet on depression have focused on single nutrients or foods only (12–15). But, of course, different populations across different settings consume different combination of food groups (16,17). The consumption of different food groups may synergistically interact with each other resulting in larger effect sizes that can modify the progression of depression, compared to the consumption of individual food group whose effect size maybe too small to be clinically meaningful (17). Thus, examining the consumption of different food groups (‘dietary diversity’), and its relationship with depression outcomes may provide information on more realistic and effective intervention targets than from evaluation of single nutrients alone.

Previous studies on the association between dietary diversity and depression have been confined to high-income countries with mixed results. In high income countries, some previous research has found that dietary diversity is negatively associated with the risk of depression (18–24). Yet, other high-income country studies showed no association between dietary diversity and risk of depression (25–28). The inconsistent findings between studies in high-income countries and contextual differences in dietary intake and patterns, limit the generalizability of study findings to low-income settings, particularly in sub-Saharan Africa. While limited, emerging scientific evidence in sub-Saharan Africa support the association between dietary diversity and maternal depression (29,30), and dietary self-care adherence and risk of depression among people living with diabetes (15). Because these studies focus on specific population sub-groups, a knowledge gap remains in understanding this relationship in rural western Kenya. We hypothesized that participants who reported higher dietary diversity would experience lower depressive symptoms compared to participants who reported lower dietary diversity in rural western Kenya.

## Methods

### Study Population and Setting

We conducted a cross-sectional study within two neighbouring rural communities with similar sociodemographic characteristics in Milo and Matulo wards located within Webuye sub-County of Bungoma county in rural western Kenya. Each ward had a population of approximately 15,000 residents at the time of survey. Bungoma County is a catchment area for the on-going population health initiatives under the Academic Model Providing Access to Healthcare (AMPATH). AMPATH is a consortium of North American Universities, Moi University and Moi Teaching and Referral Hospital (MTRH) in Kenya. Under AMPATH, a group-based microfinance program has been established, the Bridging Income Generation with Group Integrated Care (BIGPIC). We used purposive sampling to draw 312 participants and excluded one observation with missing dietary diversity data, resulting in analytic sample of 311 eligible study participants. These participants were obtained from a current roster of 4545 active BIGPIC microfinance members spread across the two study wards (Figure 1). Eligible study participants were required to be 18 years and older, current residents of Milo and Matulo wards for at least one year, and provided informed consent to participate in the study.

**Figure 1:**
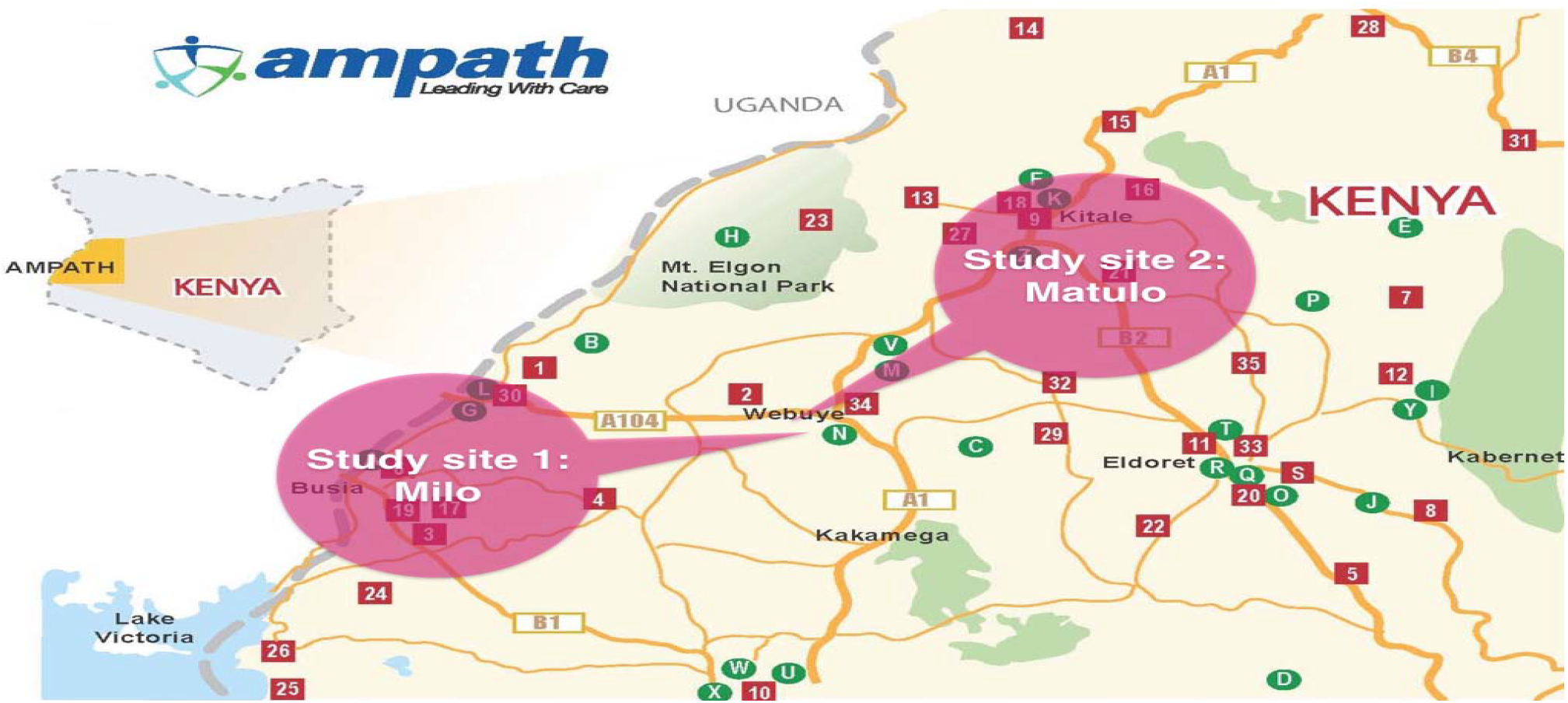
Map of BIGPIC/AMPATH program coverage in western Kenya, with study sites in Milo and Matulo Communities. Adapted from: http://ampath-uoft.ca/about-us/activities/map/

### Data Collection

We collected primary data between June and August 2025. Prior to data collection activities, three local fieldworkers were recruited and trained on the study background, objectives, research ethics, and data collection procedures using REDCap(31) software installed on tablets. The electronic questionnaire captured information on sociodemographic characteristics, dietary diversity, and depressive symptoms. To ensure cultural appropriateness and ease of comprehension, the questionnaire was translated into the local Kibukusu language. A pilot test (n=5) of the data collection instrument was conducted to assess face validity and correct for any errors that emerged.

### Ethical Consideration

We sought two ethical approvals from the Indiana University Institutional Review Board (#25993) and Moi University Institutional Research and Ethics Committee (#0005080). Permission to conduct the study was granted by the Kenya National Commission for Science, Technology, and Innovation (#P/25/4175009). Interviews were administered during weekly group meetings in private and comfortable settings after participants received information about the study and provided informed consent.

### Key Variables

The independent variable of interest (exposure) was dietary diversity. To measure dietary diversity, we assessed the total number of food groups consumed in the participants’ households during the reference recall period of 24 hours using a validated dietary diversity scale in Kenya. The validated dietary diversity scale has all the five food groups intended for the general population. Participants were asked, “yesterday did you and your family eat any of the following food groups? starch (e.g. Maize ugali, maize porridge, rice, bread, chapati, injera, pasta, or noodles); fruits (e.g. Pawpaw, mango, passion fruit, or matunda ya damu); vegetables (e.g. collard greens, Ethiopian kale, spinach, nightshade, amaranth, saget, or cowpea leaves); animal proteins (e.g. goat meat, beef, minced beef, mutton, liver or matumbo), and pulses, nuts or seeds. The greater the number of food groups consumed, the higher the likelihood that participants received all the necessary nutrients required for a healthy diet. Using a dietary diversity cut-off of five food groups, we created a binary variable where dietary diversity was equal to 1 if participants consumed all the five food groups, and 0 if participants consumed less than the five food groups. The scale was appropriate to measure dietary diversity because it uses sentinel food approach that counts the number of context-specific food groups regularly consumed by the vast majority of Kenyan population (32). The validated scale has also been used to measure dietary diversity by capturing context specific food groups in other similar setting.(33)

The dependent variable of interest (outcome) was depressive symptoms, measured with the Centre of Epidemiologic Studies Depression Scale (CED-S) (34). The CED-S scale had been validated and used previously to evaluate depression severity in a similar setting (35). Participants were asked 20 questions that describe “How often have you been bothered by any of the following problems during the past one week?” The response for each problem was rated on a 4-point Likert Scale with the options “ 0 = rarely or none of the time, “ “1 = sometime or little of the time,” “2 = Occasionally or moderate amount of the time,” “ 3 = most or all of the time.” The participants’ responses were coded as 0, 1, 2, 3 from “rarely or none of the time” to “most or all of the time,” giving a cumulative score ranging from 0 to 53 which reflected the severity of depression over the last seven days. Higher scores indicated higher depressive symptoms.

### Covariates

We used a set of covariates to characterize the study sample and adjust for confounding. Specifically, age was self-reported as the number of years from the date of birth, sex was self-reported as either male or female, educational attainment was self-reported as “some primary,” “primary,” “secondary,” and “post-secondary”, household size was self-reported as the number of people in the household during the study period, marital status was self-reported as “never married,” “currently married,” and “divorced or separated”, work status self-reported as either worked outside home for income or not in the past 30 days. We calculated a weighted wealth index derived from 20 household items and categorized participants into quartiles as per the standard procedures of the 2022 Kenya Demographic Health Survey.(36)

## Statistical Analysis

We fitted linear regression models to estimate the association between high dietary diversity and continuous depression scores. We further explored the potential differences in associations between dietary diversity and depressive symptoms in low- and high-wealth status backgrounds, by including an interaction term between dietary diversity and wealth status. We conducted a secondary analysis using quantile regression to estimate the associations across the distribution of depressive symptoms. We reported the point estimates together with their respective 95% confidence intervals and p-values. Because the distribution of scores for depressive symptoms was highly skewed and bounded (0-53), we conducted a sensitivity analysis using ordinal logistic regression model. The ordinal logistic regression approach treats the scores for depressive symptoms as ordered categorical outcome variable, and reduces the influence of extreme outliers to allow for flexible assessment of the association between dietary diversity and ordered scores for depressive symptoms. We then compared the ordinal logistic regression results with those from the linear regression model to determine if there were changes in the magnitude and direction of effect estimates.

We considered the following covariates as potential confounders: wealth, age, education attainment, marital status, and household size. Our primary adjustment set included only the covariates that were statistically associated with dietary diversity: wealth and education. Given that this covariate set may not sufficiently capture all measured confounders, we compared these adjusted results to models adjusted for the full covariate set. To assess the sensitivity of our results to any unmeasured confounding, we computed E-values for the relationship between dietary diversity and depressive symptoms. The E-value on a risk ratio scale measures the minimal degree of association that unmeasured confounder would have on the exposure and outcome to completely explain away the observed association. We examined how the magnitude of E-value relates to the observed association for the measured covariates included in the model. All statistical analyses were conducted using R statistical software (version 4.4.2, Vienna, Austria).

## Results

### Sample Characteristics

We interviewed 311 eligible participants from a current roster of 4545 current BIGPIC participants during the study period. From the overall sample, most participants were women (87%), currently married (85%), and did not work outside their homes in the last 30 days for income (78%). The mean household size was 5 members (SD=2.28). Most participants age ranged between 30 and 59 years with a median age of 40 years. Compared to participants with lower dietary diversity, participants with higher dietary diversity were more likely to have attained secondary and post-secondary education, and be in the highest wealth quantile (Table 1).

**Table 1:** Study population characteristics in the full sample and by household dietary diversity exposure status (311)

|  |  |  |  |  |
| --- | --- | --- | --- | --- |
| **Household Dietary Diversity Status** |  |  |  |  |
| **Sociodemographic characteristics** | **Overall sample**<br>**(100%)** | **High Dietary**<br>**Diversity**<br>**N = 168** | **Low Dietary**<br>**Diversity**<br>**N=143** | **p-value** |
| **Sex** |  |  |  | 0.71 |
| Male | 40 (13%) | 20 (12%) | 20 (14%) |  |
| Female | 271 (87%) | 148 (88%) | 123 (86%) |  |
| **Age Category** |  |  |  |  |
| <30 | 32 (10%) | 13 (8%) | 19 (13%) | 0.35 |
| 30-39 | 105 (34%) | 55 (33%) | 50 (35%) |  |
| 40-59 | 125 (40%) | 72 (43%) | 53 (37%) |  |
| 60 + | 49 (16%) | 28 (16%) | 21 (15%) |  |
| **Marital status** |  |  |  | 0.9 |
| Never married | 46 (15%) | 25 (15%) | 21 (15%) |  |
| Currently married | 263 (85%) | 143 (85%) | 120 (85%) |  |
| Missing | 2 | 0 | 2 |  |
| **Education status** |  |  |  | **0.008** |
| None/Some Primary | 67 (21%) | 24 (14%) | 42 (29%) |  |
| Primary | 146 (46%) | 82 (49%) | 64 (44%) |  |
| Secondary | 67 (21%) | 41 (24%) | 26 (18%) |  |
| Post-secondary | 32 (10%) | 21 (13%) | 11 (8%) |  |
| **Worked outside home**<br>**(last 30 days)** |  |  |  | 0.2 |
| Yes | 69 (22%) | 38 (23%) | 31 (22%) |  |
| No | 243 (78%) | 130 (77%) | 112 (78%) |  |
| **Household size**<br>**[Mean (SD)]** | 5 | 5 (2.28) | 5 (2.28) | 0.3 |
| **Wealth quartile<sup>a</sup>** |  |  |  | **0.004** |
| Q1(poorest) | 84 (27%) | 36 (22%) | 48 (31%) |  |
| Q2 | 69 (22%) | 34 (17%) | 35 (28%) |  |
| Q3 | 81 (26%) | 45 (31%) | 36 (21%) |  |
| Q4 (wealthiest) | 70 (22%) | 50 (27%) | 20 (18%) |  |
| Missing | 7 | 3 | 4 |  |

*\*p-value reported for chi-square test of categorical variables and t-test for continuous variables.*
*Wealth quartile calculated by summing the values of 20 household items owned by each participant household.*

### Dietary Diversity and Depressive Symptoms

In the unadjusted linear model, higher dietary diversity was associated with lower depressive symptoms [unadjusted β (95% CI): -4.75 (-7.83, -1.67)]. In the model adjusted for wealth and education, we found that higher dietary diversity was associated with over 3 points lower depressive symptom scores [adjusted β (95% CI): -3.49 (-6.62, -0.38)]. The model with the expanded set of covariates (age, household size, wealth, education attainment, work status, and marital status) produced a point estimate [adjusted β (95% CI): -3.47 (-6.65, -0.28)], that was similar in magnitude and direction to the model adjusted for only wealth and education.

The interaction between dietary diversity and wealth status suggested differences in the magnitude and direction of the relationship between low- and high-wealth status [Wald p-value for interaction term =0.0003]. In the wealth-stratified model adjusted for education, the association between dietary diversity and depressive symptoms was stronger in those with low-wealth backgrounds [adjusted β (95% CI): -6.00 (-10.56, -1.42)] relative to those with high-wealth backgrounds [adjusted β (95% CI): -0.53 (-4.76, 3.68); Wald p-value for interaction term =0.0003, Table 2).

**Table 2:**
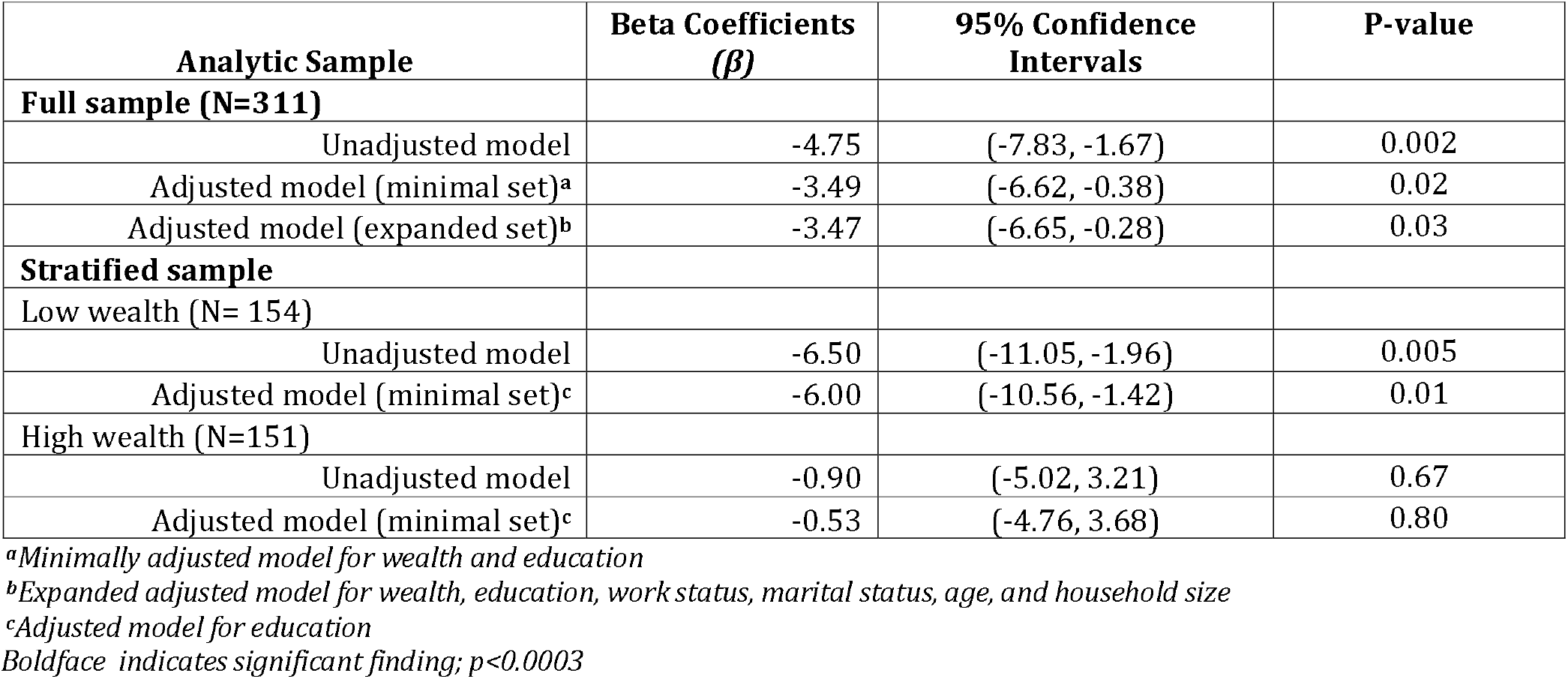
Linear regression estimates for the association between household dietary diversity and self-reported depression scores overall, and stratified by wealth status.

To further explore how the association varied across the distribution scores for depressive symptoms, we conducted a secondary analysis using quantile regression. Across all quantiles, the effect estimates consistently indicated lower scores for depressive symptoms among participants with higher dietary diversity, showing a negative association in both unadjusted and adjusted models. In the unadjusted model, the associations were strongest, with larger effect sizes observed across 75^th^ (Q75: β = -7.56; 95% CI: -12.82, -2.27), and 90^th^ (Q90: β = - 6.01; 95% CI: -12.43, 0.43) quantiles, compared to effect sizes observed across lower quantiles for 10^th^ (Q10: β = -1.0; 95% CI: −2.74, 0.74) and 90^th^ (Q90: β = -1.0; 95% CI: -5.12, 3.12) quantiles. After adjustment for wealth and education, the magnitude of the associations was substantially attenuated, with the 95% confidence intervals crossing the null at the 10^th^ (Q10: β = -0.59; 95% CI: -2.46, 1.28), 25^th^ [Q25: β = -0.82; 95% CI: -4.14, 2.50), 75^th^ (Q75: β = −4.00; 95% CI: −10.13, 2.13), and the 90^th^ (Q90: β = −1.59; 95% CI: −7.43, 4.25). Despite this attenuation, the direction of the association remained consistently negative across higher quantiles, suggesting a potential protective pattern, albeit with reduced precision of the estimates. Overall, these findings indicate that although covariate adjustment weakened the statistical precision of the associations, the general increasing trend of the effect sizes for the observed association between higher dietary diversity and lower depressive symptom scores persisted, particularly among individuals with more severe depressive symptoms. (Table 3).

**Table 3:** Quantile regression estimates for the association between dietary diversity and self-reported depression scores across various quantiles (n=311)

Boldface indicates significant finding; $p < 0.0003$
|  |  |  |  |  |
| --- | --- | --- | --- | --- |
| **Quantile** | **Unadjusted Model** |  | **Adjusted Model** |  |
| **Q10** | **$\beta$ Coefficient (95% CI)** | **p-value** | **$\beta$ Coefficient (95% CI)** | **p-value** |
| Low Dietary Diversity | Ref. | - | Ref. | - |
| High Dietary Diversity | -1.0(-2.74, 0.74) | 0.26 | -0.59(-2.46, 1.28) | 0.54 |
| **Q25** |  |  |  |  |
| Low Dietary Diversity | Ref. | - | Ref. | - |
| High Dietary Diversity | -1.0(-5.12, 3.12) | 0.64 | -0.82(-4.14, 2.50) | 0.63 |
| **Q50** |  |  |  |  |
| Low Dietary Diversity | Ref | - | - | - |
| High Dietary Diversity | -6.28(-11.65, -0.91) | **0.02** | -4.00(-10.13, 2.13) | 0.11 |
| **Q75** |  |  |  |  |
| Low Dietary Diversity | Ref | - | - | - |
| High Dietary Diversity | -7.56(-12.85, -2.27) | **0.005** | -4.00(-9.49, 3.49) | 0.20 |
| **Q90** |  |  |  |  |
| Low Dietary Diversity | Ref | - | Ref. | - |
| High Dietary Diversity | -6.01(-12.43, 0.43) | **0.06** | -1.59(-7.43, 4.25) | 0.59 |

Covariates in the minimal adjustment set were education, and wealth.
95% CI: Confidence Interval.
Boldface indicates significant finding; $p < 0.05$

To explore how much unmeasured confounding might have influenced our results, we calculated the E-value for the association between dietary diversity and depressive symptoms. The E-value for the adjusted effect estimate was 1.82, meaning that an unmeasured confounding factor would need to be fairly strongly related to both dietary diversity and depressive symptoms by a risk ratio of at least 1.82 each, to completely explain away the association we observed. The lower bound of the 95% confidence interval corresponded to an E-value of 1.16, indicating that unmeasured confounder with a more modest association could move the 95% confidence interval to overlap the null. Our sensitivity analysis results showed that the E-value for the point estimate was 1.82 (95% CI lower bound:1.16), suggesting that unmeasured confounder would need to have an association with both dietary diversity and depressive symptoms that is larger in magnitude than most of the measured covariates included in the model: age, work status, household size, wealth status and education which were apriori considered among the strongest confounders to fully explain away the observed association. Overall, these results indicate that the observed association was reasonably robust to unmeasured confounding (Table 4)

**Table 4:** Linear regression estimates for covariates and e-value for the dietary diversity exposure.

|  |  |  |  |
| --- | --- | --- | --- |
| **Exposure variable:** | **$\beta$ (95% CI)** | **E-value** | **Lower 95% CI** |
| **Dietary diversity** | -3.47 (-6.65, -0.28) | 1.82 | 1.16 |
| **Covariate adjustment set** |  |  |  |
| Work status | -1.74 (-5.44, 1.96) |  |  |
| Wealth status | -6.39 (-11.26, -2.78) |  |  |
| Age | 0.05 ( 0.99, 1.01) |  |  |
| Household size | 0.01 (-0.68, 0.71) |  |  |
| Marital status | 2.50 (-2.09, 7.08) |  |  |
| Education | 5.51 (-7.88, 1.89) |  |  |

In another sensitivity analysis, results from an ordinal logistic regression model showed that higher dietary was associated with lower cumulative odds of more severe depressive symptoms, (AOR=0.64; 95% CI: 0.41, 1.00), suggesting that, across thresholds, participants with higher dietary diversity had approximately 36% lower cumulative odds of being in a worse depression category, after adjusting for covariates (Table 5). These results were consistent with our main results from linear regression model, indicating robustness of our findings to alternative model specifications.

**Table 5:** Sensitivity analysis using ordinal logistic regression to estimate the association between dietary diversity and ordered categorical depression outcome variable.

|  |  |  |  |
| --- | --- | --- | --- |
|  | **Adjusted Odds Ratio (AOR)** | **95% Confidence Interval** | **p-value** |
| Dietary diversity categories |  |  |  |
| Low | Ref. |  |  |
| High | 0.64 | 0.41, 1.00 | 0.05 |

*Model adjusted for age, household size, wealth quantile, education, and work status*

## Discussion

We examined the relationship between higher dietary diversity and depressive symptoms in rural western Kenya. We found that higher dietary diversity was associated with lower depressive symptoms, particularly among participants from low-wealth backgrounds. We also found that the strength of association differed across the distribution of depression scores: Higher dietary diversity tended to be more strongly associated with lower depression scores in individuals with more severe symptoms. These findings highlight the potential of improving dietary diversity as a strategy to address mental health, particularly among individuals from lower-wealth backgrounds with worse depressive symptoms in low-income settings.

Our main findings align with much of the evidence base in high-income countries, supporting the conclusion that higher dietary diversity is associated with lower depressive symptoms (16,18–21,24,37–40). However, the strong associations we observed between dietary diversity and lower depressive symptoms among individuals from lower-wealth backgrounds was unexpected. A previous study found that food security was more strongly related to better psychological outcomes in high-income and education households compared with low-income and education households (41). The inconsistent findings may be explained by differences in how socioeconomic status was measured between our study and prior work. In our study, wealth status was examined as an effect measure modifier of the association between dietary diversity and depressive symptoms, whereas the previous study used income and educational attainment as effect measure modifiers in assessing the relationship between food insecurity and depression outcome. These differences in the operationalization of socioeconomic status may have contributed to the observed discrepancies in results.

There are plausible pathways through which higher dietary diversity may lead to a reduction in depressive symptoms. First, the consistent consumption of diverse food groups, specifically vegetables, cereals and proteins may reduce oxidative stress and inhibit brain signalling which could lead to reduced depressive symptoms among adults (21). Specifically, the protective role of fibres found abundantly in vegetables and cereals, and fruits may prevent the onset of depressive symptoms by modifying the composition and activity of microbiota in the digestive system through the gut-brain axis and metabolic mechanisms involved in depression (42,43). Secondly, adequate dietary diversity may protect against depression by providing broader and more sufficient nutrients, which can lower dietary inflammatory potential, support neurotransmitter synthesis, stabilise glycaemic control and reduce psychosocial stressors associated with the occurrence of depressive symptoms (44). Finally, and through psychosocial and socioeconomic pathways, higher dietary diversity may be protective against risk of depression by reducing food insecurity and economic stress which are established risk factors for depression (45), particularly among the socioeconomically disadvantaged population sub-groups.

Some aspects of our current study require careful interpretation of results. Because we measured depression and dietary diversity at the same time point, we were unable to establish the temporal ordering between the two variables. Consequently, reverse causality cannot be ruled out. Future prospective studies should clarify the directionality of the observed association in these settings. Social desirability bias in the measurement of depression was likely, as some participants may have perceived certain questions as sensitive or felt compelled to give socially acceptable responses about their symptoms. To minimize this potential misclassification bias, we translated the depression scale into the local language for comprehension and cultural appropriateness. In addition, we included an introductory statement in the questionnaires to normalize experiences of depressive symptoms and encourage honest reporting.

Adequate control of confounding was a major concern in our study. We attempted to address this concern by controlling for wealth and education in the primary adjusted model, which were statistically associated with both dietary diversity and depressive symptoms. We then compared the magnitude of the effect estimates to a model that included all the measured covariates The results suggested no statistically significant differences in either the magnitude or direction of the point estimates between the primary and fully adjusted models. The consistent results indicate that the observed association was not materially influenced by the inclusion of additional covariates, supporting the robustness and stability of our findings. We also computed E-values to assess for the minimum strength that an unmeasured confounder would need to have to completely explain away the observed association. The E-value analysis indicated that the observed association was reasonably robust to unmeasured confounding, thus strengthening the confidence in our findings. However, the possibility of uncontrolled confounding remains and further research using more robust approaches (e.g., natural experiments, randomization) would help strengthen inference.

The generalizability of our results is limited. Our study sample was participants in a group-based microfinance program in rural western Kenya so the generalizability of our findings to people in other settings and those not involved in group-based microfinance is untested. However, the plausible pathways through which we may have observed the association between dietary diversity and depressive symptoms (biological and socioeconomic pathways) may hold true across populations, supporting cautious generalizability of our study findings to other similar resource-poor contexts. Nevertheless, our study adds to the existing literature, the evidence on the association between higher dietary diversity and lower depressive symptoms from rural western Kenya where depression and undernutrition often co-occur.

## Conclusion

We found that high dietary diversity was associated with lower depression scores in rural western Kenya. These findings provide evidence that greater dietary diversity may offer some protection against depressive symptoms, consistent with patterns reported in studies from higher-income countries. Future research in resource-limited settings should prioritize longitudinal and quasi-experimental designs to strengthen causal inference and identify the mechanisms through which dietary diversity may influence depression.

## Data Availability

The primary data for this study is not publicly available due to sensitive information about study participants. However, they may be made available upon reasonable request to the corresponding author.

## References

1. World Health Organization. Depression. World Health Organization. 2025. https://www.who.int/news-room/fact-sheets/detail/depression

2. Global-Minded. Depression in Kenya. Global-Minded. 2023. https://www.globallyminded.org/home/depression-in-kenya/

3. Marangu, E., Mansouri, F., Sands, N., Ndetei, D., Muriithi, P., Wynter, K., & Rawson, H. (2021). Assessing mental health literacy of primary health care workers in Kenya: a cross-sectional survey. International journal of mental health systems, 15(1), 55. 10.1186/s13033-021-00481-z

4. Kumar, M., Njuguna, S., Amin, N., Kanana, S., Tele, A., Karanja, M., … & Weaver, M. R. (2024). Burden and risk factors of mental and substance use disorders among adolescents and young adults in Kenya: results from the Global Burden of Disease Study 2019. EClinicalMedicine, 67. DOI: 10.1016/j.eclinm.2023.102328 External Link

5. Larsen, A., Pintye, J., Marwa, M. M., Watoyi, S., Kinuthia, J., Abuna, F., … & John-Stewart, G. (2022). Trajectories and predictors of perinatal depressive symptoms among Kenyan women: a prospective cohort study. The Lancet Psychiatry, 9(7), 555–564. DOI: 10.1016/S2215-0366(22)00110-9 External Link

6. Saveanu, R. V., & Nemeroff, C. B. (2012). Etiology of depression: genetic and environmental factors. Psychiatric clinics, 35(1), 51–71. DOI: 10.1016/j.psc.2011.12.001 External Link

7. Tiemeier, H. (2003). Biological risk factors for late life depression. European journal of epidemiology, 18(8), 745–750. 10.1023/A:1025388203548

8. Buckman, J. E., Underwood, A., Clarke, K., Saunders, R., Hollon, S. D., Fearon, P., & Pilling, S. (2018). Risk factors for relapse and recurrence of depression in adults and how they operate: A four-phase systematic review and meta-synthesis. Clinical psychology review, 64, 13–38. 10.1016/j.cpr.2018.07.005

9. Hammen, C. (2018). Risk factors for depression: an autobiographical review. Annual review of clinical psychology, 14(1), 1–28. 10.1146/annurev-clinpsy-050817-084811

10. Jacka, F. N., Pasco, J. A., Mykletun, A., Williams, L. J., Hodge, A. M., O’Reilly, S. L., … & Berk, M. (2010). Association of Western and traditional diets with depression and anxiety in women. American journal of psychiatry, 167(3), 305–311. 10.1176/appi.ajp.2009.09060881

11. Sanchez-Villegas, A., & Martínez-González, M. A. (2013). Diet, a new target to prevent depression?. BMC medicine, 11(1), 3. 10.1186/1741-7015-11-3

12. Murakami, K., & Sasaki, S. (2010). Dietary intake and depressive symptoms: a systematic review of observational studies. Molecular nutrition & food research, 54(4), 471–488. 10.1002/mnfr.200900157

13. Sanhueza, C., Ryan, L., & Foxcroft, D. R. (2013). Diet and the risk of unipolar depression in adults: systematic review of cohort studies. Journal of Human Nutrition and Dietetics, 26(1), 56–70. 10.1111/j.1365-277X.2012.01283.x

14. Ainamani, H. E., Bamwerinde, W. M., Rukundo, G. Z., Tumwesigire, S., Mfitumukiza, V., Bikaitwoha, E. M., & Tsai, A. C. (2021). Fruit and vegetable intake and mental health among family caregivers of people with dementia in Uganda. Mental Health & Prevention, 24, 200223. 10.1016/j.mhp.2021.200223

15. Duodu, P. A., Okyere, J., Akyirem, S., Waring, G., & Gillibrand, W. (2025). Association between dietary self-care adherence and depression among adults living with type 2 diabetes mellitus in Ghana: a cross-sectional study. Journal of Health, Population and Nutrition, 44(1), 210. 10.1186/s41043-025-00859-6

16. Akbaraly, T. N., Sabia, S., Shipley, M. J., Batty, G. D., & Kivimaki, M. (2013). Adherence to healthy dietary guidelines and future depressive symptoms: evidence for sex differentials in the Whitehall II study1. The American journal of clinical nutrition, 97(2), 419. 10.3945/ajcn.112.041582

17. Hu, F. B. (2002). Dietary pattern analysis: a new direction in nutritional epidemiology. Current opinion in lipidology, 13(1), 3–9.

18. Dipnall, J. F., Pasco, J. A., Meyer, D., Berk, M., Williams, L. J., Dodd, S., & Jacka, F. N. (2015). The association between dietary patterns, diabetes and depression. Journal of affective disorders, 174, 215–224. 10.1016/j.jad.2014.11.030

19. Ruusunen, A., Lehto, S. M., Mursu, J., Tolmunen, T., Tuomainen, T. P., Kauhanen, J., & Voutilainen, S. (2014). Dietary patterns are associated with the prevalence of elevated depressive symptoms and the risk of getting a hospital discharge diagnosis of depression in middle-aged or older Finnish men. Journal of affective disorders, 159, 1–6. 10.1016/j.jad.2014.01.020

20. Rahe, C., Unrath, M., & Berger, K. (2014). Dietary patterns and the risk of depression in adults: a systematic review of observational studies. European journal of nutrition, 53(4), 997–1013. 10.1007/s00394-014-0652-9

21. Lai, J. S., Hiles, S., Bisquera, A., Hure, A. J., McEvoy, M., & Attia, J. (2014). A systematic review and meta-analysis of dietary patterns and depression in community-dwelling adults. The American journal of clinical nutrition, 99(1), 181–197. 10.3945/ajcn.113.069880

22. Akbaraly, T. N., Brunner, E. J., Ferrie, J. E., Marmot, M. G., Kivimaki, M., & Singh-Manoux, A. (2009). Dietary pattern and depressive symptoms in middle age. The British Journal of Psychiatry, 195(5), 408–413. doi:10.1192/bjp.bp.108.058925

23. Hu, D., Cheng, L., & Jiang, W. (2019). Sugar-sweetened beverages consumption and the risk of depression: A meta-analysis of observational studies. Journal of affective disorders, 245, 348–355. 10.1016/j.jad.2018.11.015

24. Oliván-Blázquez, B., Aguilar-Latorre, A., Motrico, E., Gómez-Gómez, I., Zabaleta-del-Olmo, E., Couso-Viana, S., … & Magallón-Botaya, R. (2021). The relationship between adherence to the Mediterranean diet, intake of specific foods and depression in an adult population (45–75 years) in primary health care. A cross-sectional descriptive study. Nutrients, 13(8), 2724. 10.3390/nu13082724

25. Gougeon, L., Payette, H., Morais, J., Gaudreau, P., Shatenstein, B., & Gray-Donald, K. (2015). Dietary patterns and incidence of depression in a cohort of community-dwelling older Canadians. The journal of nutrition, health & aging, 19(4), 431–436. 10.1007/s12603-014-0562-9

26. Sugawara, N., Yasui-Furukori, N., Tsuchimine, S., Kaneda, A., Tsuruga, K., Iwane, K., … & Kaneko, S. (2012). No association between dietary patterns and depressive symptoms among a community-dwelling population in Japan. Annals of general psychiatry, 11(1), 24. 10.1186/1744-859X-11-24

27. Jacka, F. N., Mykletun, A., Berk, M., Bjelland, I. & Tell, G. S. (2011). The Association Between Habitual Diet Quality and the Common Mental Disorders in Community-Dwelling Adults. Psychosomatic Medicine, 73 (6), 483–490. doi: 10.1097/PSY.0b013e318222831a.

28. Chatzi, L., Melaki, V., Sarri, K., Apostolaki, I., Roumeliotaki, T., Georgiou, V., … & Kogevinas, M. (2011). Dietary patterns during pregnancy and the risk of postpartum depression: the mother–child ‘Rhea’cohort in Crete, Greece. Public health nutrition, 14(9), 1663–1670. doi:10.1017/S1368980010003629

29. Kalam, M. A., McCann, J. K., Shakil, Z., Gambari, A., Ochieng, M., & Jeong, J. (2025). Maternal mental health and child dietary diversity in rural Kenya: Findings from a pooled analysis of two baseline studies. Current Developments in Nutrition, 107497. 10.1016/j.cdnut.2025.107497

30. Madeghe, B. A., Kogi-Makau, W., Ngala, S., & Kumar, M. (2021). Nutritional factors associated with maternal depression among pregnant women in urban low-income settlements in Nairobi, Kenya. Food and Nutrition Bulletin, 42(3), 334–346. 10.1177/03795721211025123

31. Harris, P. A., Taylor, R., Minor, B. L., Elliott, V., Fernandez, M., O’Neal, L., … & REDCap Consortium. (2019). The REDCap consortium: building an international community of software platform partners. Journal of biomedical informatics, 95, 103208. 10.1016/j.jbi.2019.103208

32. Global Diet Quality Project. Diet Quality Questionnaire (DQQ) for Kenya (2021). Accessed at https://dietquality.org on March 6, 2025.

33. Ma, S., Herforth, A. W., Vogliano, C., & Zou, Z. (2022). Most commonly-consumed food items by food group, and by province, in China: implications for diet quality monitoring. Nutrients, 14(9), 1754. 10.3390/nu14091754

34. Radloff, L. S. (1977). A self-report depression scale for research in the general population. Applied psychol Measurements, 1, 385–401. 10.1177/014662167700100306

35. Natamba, B. K., Achan, J., Arbach, A., Oyok, T. O., Ghosh, S., Mehta, S., … & Young, S. L. (2014). Reliability and validity of the center for epidemiologic studies-depression scale in screening for depression among HIV-infected and-uninfected pregnant women attending antenatal services in northern Uganda: a cross-sectional study. BMC psychiatry, 14(1), 303. 10.1186/s12888-014-0303-y

36. Kenya National Bureau of Statistics. Demographic and Health Survey (2022). https://www.knbs.or.ke/reports/kdhs-2022/

37. Geldsetzer, P., Vaikath, M., Wagner, R., Rohr, J. K., Montana, L., Gómez-Olivé, F. X., … & Berkman, L. F. (2019). Depressive symptoms and their relation to age and chronic diseases among middle-aged and older adults in rural South Africa. The Journals of Gerontology: Series A, 74(6), 957–963. doi:10.1093/gerona/gly145

38. Yin, W., Löf, M., Chen, R., Hultman, C. M., Fang, F., & Sandin, S. (2021). Mediterranean diet and depression: a population-based cohort study. International Journal of Behavioral Nutrition and Physical Activity, 18(1), 153. 10.1186/s12966-021-01227-3

39. Oddo, V. M., Welke, L., McLeod, A., Pezley, L., Xia, Y., Maki, P., … & Tussing-Humphreys, L. (2022). Adherence to a Mediterranean diet is associated with lower depressive symptoms among US adults. Nutrients, 14(2), 278. 10.3390/nu14020278

40. Matta, J., Hoertel, N., Airagnes, G., Czernichow, S., Kesse-Guyot, E., Limosin, F., … & Lemogne, C. (2020). Dietary restrictions and depressive symptoms: Longitudinal results from the constances cohort. Nutrients, 12(9), 2700. 10.3390/nu12092700

41. Militao, E. M., Uthman, O. A., Salvador, E. M., Vinberg, S., & Macassa, G. (2024). Association between socioeconomic position of the household head, food insecurity and psychological health: an application of propensity score matching. BMC Public Health, 24(1), 2590. 10.1186/s12889-024-20153-0

42. Liu, X., Cao, S., & Zhang, X. (2015). Modulation of gut microbiota–brain axis by probiotics, prebiotics, and diet. Journal of agricultural and food chemistry, 63(36), 7885–7895.

43. Sanchez-Villegas, A., Zazpe, I., Santiago, S., Perez-Cornago, A., Martinez-Gonzalez, M. A., & Lahortiga-Ramos, F. (2018). Added sugars and sugar-sweetened beverage consumption, dietary carbohydrate index and depression risk in the Seguimiento Universidad de Navarra (SUN) Project. British Journal of Nutrition, 119(2), 211–221. doi:10.1017/S0007114517003361

44. Jacka, F. N., O’Neil, A., Opie, R., Itsiopoulos, C., Cotton, S., Mohebbi, M., … & Berk, M. (2017). A randomised controlled trial of dietary improvement for adults with major depression (the ‘SMILES’trial). BMC medicine, 15(1), 23. DOI: 10.1186/s12916-017-0791-y

45. Pengpid, S., & Peltzer, K. (2024). Dietary diversity among women with depressive and generalized anxiety symptoms in Nepal. Scientific Reports, 14(1), 17688. DOI: 10.1038/s41598-024-68346-2

